# An example of too much too soon? A retrospective review of Caesarean Sections performed in the first stage of labour in Kenya

**DOI:** 10.1101/2023.07.30.23293379

**Authors:** Helen Allott, Fiona Dickinson, Stephen Karangau, Michael Oduor, Nassir Shaaban, Evans Ogoti, Sheila Sawe, Ephraim Ochola, Charles Ameh

## Abstract

**Objective:** In Kenya, decisions to perform CS are frequently made by unsupported non-specialist doctors, sometimes resulting in sub-optimal decision-making and inappropriate surgery. This study assesses decision-making in CS performed in the first stage of labour by a retrospective case review.

**Methods:** A panel of expert obstetricians reviewed case-notes randomly selected from a series obtained from seven Kenyan referral hospitals, then discussed in pairs and as a group where opinions differed.

**Results:** Of the 87 case-notes reviewed consensus was achieved in 94.3% cases. In 41.3% cases, CS was considered appropriate by all assessors, including 8.9% cases where the CS was necessary but performed too late. The decision to delivery interval was 2 hours or longer in 65.8% cases, including 18 cases done for non-reassuring fetal status. In 10.3% it was considered that further reassessment should have occurred.

In 9.2% the reviewers concluded that the CS was done too soon and alternative measures could have been taken. Insufficient information was available in the records to make a full assessment in 21.8% of cases and in 11.5% the CS was considered to be inappropriate.

**Conclusions:** This review suggests a need for improved support for decision-making, improved record-keeping and more timely surgery when indicated.

**Synopsis:** At least 11.5% of Caesarean Sections in the first stage of labour could be avoided if decision-makers had access to support from more experienced practitioners.

## Introduction

Caesarean Section was reported as an intervention to relieve obstructed labour in Africa as early as 1879 when Robert Felkin observed the procedure in Uganda performed on a woman partially anaesthetised with banana wine [1]. On that occasion both mother and child survived, although mortality from the procedure at that time was reported to be high. Since then, there have been many improvements in surgical safety and there is no doubt that appropriate use of CS is life-saving for both mothers and babies. However, CS is still associated with both short and long-term consequences, with CS-associated mortality reported in a systematic review as 10.9 per 1000 procedures in sub-Saharan Africa [2]. In the longer term, CS is associated with significant maternal risks in future pregnancy, including uterine rupture and abnormal placentation, with attendant risks of catastrophic haemorrhage [3, 4]. For the neonate, short-term risks of CS include altered immune development, an increased likelihood of allergy, atopy, and asthma, and reduced intestinal gut microbiome diversity [5]. Given these risks, it is important that CS usage is restricted to those cases with appropriate clinical indications.

Moreover, considering the limited surgical capacity of many health facilities, especially in low-income countries, conducting non-essential surgery may cause delays for those cases with clear indications. This knock-on effect may contribute to poor maternal and neonatal outcomes for those waiting in a queue for theatre.

Despite all the associated risks, there has nevertheless been a rise in rates of CS across the globe, with an estimated overall rate of 21.1% in 2015, equating to almost 30 million Caesarean births [6]. This rise has fired the debate regarding the establishment of a recommended rate for birth by CS. A systematic review of ecological studies determined an association between rising caesarean section rates and a decrease in both maternal and newborn mortality up to a ceiling varying between 9% and 16%, after which there were no further improvements in mortality as rates increased further [7]. A large ecological study including 159 countries, concluded that CS rates greater than 10% may not be significantly correlated with changes in maternal and neonatal mortality [8]. In a statement on CS rates in 2015, the WHO concluded that despite socioeconomic factors explaining a substantial part of the crude association between CS rate and mortality, nevertheless, at CS rates up to 10% there was an inverse association between CS rates and maternal and newborn mortality [9]. An association between stillbirth or morbidity outcomes and CS rates could not be determined due to the lack of data at the population level. The WHO has stated that, rather than striving to achieve a recommended CS rate, every effort should be made to provide caesarean sections to women in need [9].

In many low-income settings, medical staff who provide care for women giving birth lack specialist training or support, yet they are required to make complex decisions regarding surgical interventions. This has the potential to result in the performance of some unnecessary CSs, and equally a failure to prioritise some cases where CS should have been performed earlier in labour. Moreover, in settings with limited capacity, the performance of unnecessary CS may lead to extended delays for indicated CS, with deleterious consequences for women and babies. Solutions to the joint issues of unnecessary CS and CS performed too late are likely to be multifactorial. Amongst other strategies, the WHO has recommended that mandatory second opinions should be sought before performance of CS, and the performance of facility audit of CS with feedback. In some locations obtaining a second opinion may not be possible, leaving doctors to make a sometimes-complex management choice without the opportunity for discussion.

Expert case review is an established method utilised in undertaking confidential enquiry into maternal deaths. In such reviews experts make judgements as to whether clinical management was appropriate or, if not, whether different management could have resulted in a better outcome [10].

The aim of this study is to assess decision-making in CS performed in the first stage of labour in Kenya by a retrospective case review.

## Methods

### Study design

A retrospective expert review of randomly selected records of women who gave birth by CS in the first stage of labour in Kenyan county and sub-county hospitals, was undertaken.

### Study setting

Case notes were drawn from hospitals which were part of a United Kingdom Foreign, Commonwealth & Development Office funded, Maternal and Newborn Health Programme implemented in five of the 47 counties across Kenya. Seven high volume facilities were selected from the 25 healthcare facilities included in the programme. (High volume was defined as conducting more than 130 births per month). All included healthcare facilities were either county level or sub-county hospitals, functioning at a Comprehensive Emergency Obstetric and Newborn Care (CEmOC) level, and all accepted cases referred from other facilities.

### Ethical considerations

Ethical approval was sought prospectively and obtained from the LSTM Research Ethics Committee (ref: 21-041) and the Moi University Institutional Research Ethics Committee (ref: IREC/2021/115). Personal identifiable information concerning staff and patients was electronically redacted to preserve confidentiality and reduce the possibility of any reviewer bias.

### Sample size and data collection

A random selection of case-notes were collected for the period June to November 2020, to avoid the restrictions surrounding the initial onset of the Coronavirus pandemic and a healthcare providers strike which occurred in December 2020. CS case-notes were selected using a systematic random sampling technique, using either the Kenya Health Information System, where possible, or the hospital maternity registers, as a sampling frame.

From a total of 826 randomly selected CS case-notes retrieved, 507 (61.3%) were initially designated as eligible for this study, having been carried out in the first stage of labour. 90 cases were selected randomly from the 507 for detailed analysis, but three were excluded after it was determined by initial review that the CS had not been performed in the first stage of labour, leaving a sample size of 87. The choice of sample size was pragmatic, based upon the time that reviewers were able to commit to the study. However, for the sample size used, with a 95% confidence interval, the margin of error would not exceed 9.6%, with a confidence interval within 19.2%.

Identification, retrieval and anonymisation of medical records of included cases were carried out through the Kenya CEMD system [11]. All data collected was managed on Microsoft Excel Version 16.71 (23031200) and statistics were calculated manually.

The reviewers are all experienced obstetricians and none of them work in the hospitals included in the study. Six reviewers came from Kenya and one from the UK but with experience of working in LMICs including Kenya.

### Analysis

Reviewers extracted the following information from each case record; referral status, parity, history of previous CS, number of fetuses, presentation of the fetus, birthweight, Apgar scores at 1 and 5 minutes, newborn outcome and complications, maternal outcome and complications, stated reason for the CS, cervical dilation at decision, position of the presenting part, descent in the maternal abdomen, contraction frequency and the decision to delivery interval, into a pre-designed spreadsheet.

Based upon their review of the available information, reviewers then categorised the CS according to the following criteria: 1) CS was considered to have been necessary and appropriate, 2) CS was done too soon and alternative measures could been tried to progress labour, 3) Prolonged decision to delivery interval and a further assessment should have been undertaken to ascertain if further progress made, 4) CS necessary but done too late, 5) Insufficient information available to make a judgement as to whether CS appropriate and 6) Inappropriate. Categories 1 and 4 were judged as appropriate CS.

Each case was independently reviewed by two members of the panel and a conclusion drawn as to the potential appropriateness of the procedure. Where any discrepancies of opinion were identified between panellists following the initial review, cases were discussed by the group as a whole and a consensus formed wherever possible.

Data concerning maternal and neonatal outcomes and decision to delivery intervals were analysed as simple descriptive statistics.

## Results

Overall, 36 (41.3%) CS were considered appropriate (categories 1 and 4), including 7 (8.9%) (category 4) where it was judged that the decision should have been made earlier (Table 1).

**Table 1.**
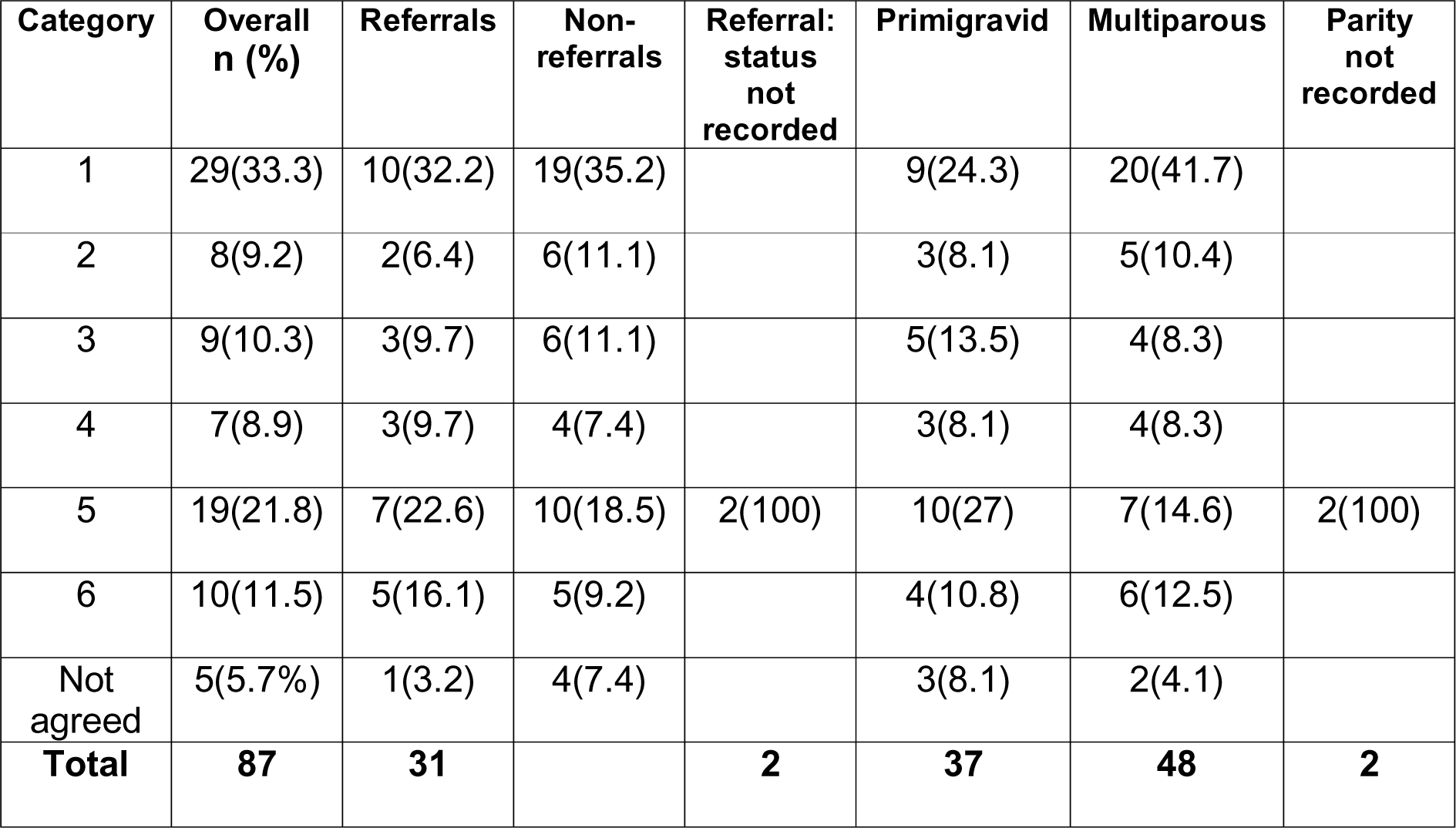
Classification and characteristics of included cases.

| Category | Overall n (%) | Referrals | Non-referrals | Referral: status not recorded | Primigravid | Multiparous | Parity not recorded |
| --- | --- | --- | --- | --- | --- | --- | --- |
| 1 | 29(33.3) | 10(32.2) | 19(35.2) |  | 9(24.3) | 20(41.7) |  |
| 2 | 8(9.2) | 2(6.4) | 6(11.1) |  | 3(8.1) | 5(10.4) |  |
| 3 | 9(10.3) | 3(9.7) | 6(11.1) |  | 5(13.5) | 4(8.3) |  |
| 4 | 7(8.9) | 3(9.7) | 4(7.4) |  | 3(8.1) | 4(8.3) |  |
| 5 | 19(21.8) | 7(22.6) | 10(18.5) | 2(100) | 10(27) | 7(14.6) | 2(100) |
| 6 | 10(11.5) | 5(16.1) | 5(9.2) |  | 4(10.8) | 6(12.5) |  |
| Not agreed | 5(5.7%) | 1(3.2) | 4(7.4) |  | 3(8.1) | 2(4.1) |  |
| **Total** | **87** | **31** |  | **2** | **37** | **48** | **2** |

In eight (9.2%) cases (category 2), the expert review concluded that a decision to perform CS had been made too soon, and alternative measures, for example, augmentation of labour, might have resulted in avoidance of CS. In nine (10.3%) cases with a prolonged decision to delivery interval (category 3), it was considered that, had a further assessment been performed prior to the CS, findings may have changed, rendering the CS potentially unnecessary. In 19 (21.8%) cases (category 5) the quality of the records was such that insufficient information was available for the experts to reach a conclusion as to the necessity of the CS and in 10 (11.5%) they concluded that there had been no valid indication for the CS (category 6). In 5 (5.7%) cases the experts were unable to reach a consensus. The common factor in these cases was the passage of meconium in utero.

When parity was taken into consideration, 12 (32.4%) of primigravid women had a CS considered appropriate, compared to 24 (50%) of multiparous women. However, when women presenting in labour who had two or more previous CS were excluded, 16(33.3%) of multiparous women had appropriate CS.

31 (35.6%) cases were accepted as referrals from other health facilities and 54 (62.1%) were admitted directly from home. In two cases the source was not determined. There was no difference to the proportion of cases judged appropriate whether cases had been referred from other facilities or admitted directly from home.

The decision to delivery interval (DDI) was identified in 79 (90.8%) cases (Figure 1), of which in 52 (65.8%) cases this was two hours or longer. Of the 25 cases where the primary reason for the CS was given as non-reassuring fetal status, in 5 (20%) the DDI was two hours or more, and this was associated with poor outcomes.

**Figure 1:** Decision to delivery interval (hours)

Of the reasons stated in the records for the CS, the most frequent were “non-reassuring fetal status” (24 cases, 28%) and “poor progress” (13 cases, 15%). In 33 of the 36 cases that the assessors considered the CS was definitely indicated, they agreed with the stated reason for the CS. In three cases they felt the CS was indicated, but not for the reasons stated in the records. This included two cases where the stated reason was failure to progress in labour and one of non-reassuring fetal status.

The most frequently missing parameters relating to decision-making documented in the medical records were the frequency and duration of the contractions (75 cases, 86%), the position of the presenting part (72 cases, 83%) and the abdominal descent of the presenting part (41 cases, 47%). Inconsistent documentation regarding the fetal heart rate and pattern was a universal finding.

Using data provided by each of the seven units, the midwifery caseload was compared to the percentage of births performed by CS (Table 2). The caseload was calculated by dividing the number of births reported per annum at the time of the study by the number of full-time equivalent midwives employed. This provides an indication as to the pressure of work experienced by the midwives.

**Table 2.**
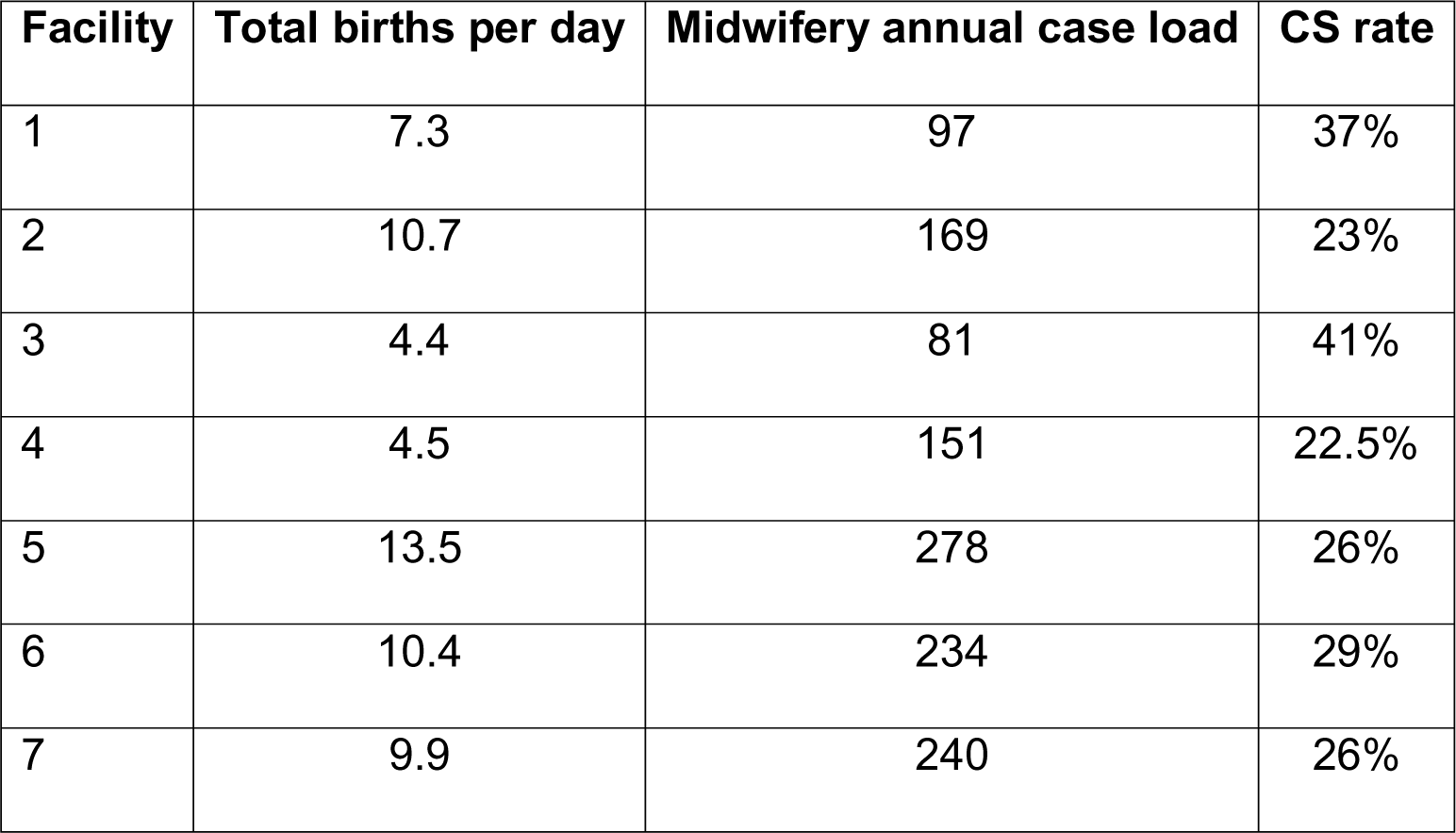
Midwifery caseload and CS rate.

| Facility | Total births per day | Midwifery annual case load | CS rate |
| --- | --- | --- | --- |
| 1 | 7.3 | 97 | 37% |
| 2 | 10.7 | 169 | 23% |
| 3 | 4.4 | 81 | 41% |
| 4 | 4.5 | 151 | 22.5% |
| 5 | 13.5 | 278 | 26% |
| 6 | 10.4 | 234 | 29% |
| 7 | 9.9 | 240 | 26% |

### Patient outcomes

Eight babies out of the 87 where Apgar Scores were recorded had scores less than 7 at 5 minutes, including two that were freshly stillborn, and there were six admissions to a newborn care unit. Three mothers experienced a post-partum haemorrhage and there was one case of bladder injury. It was not possible to assess whether there were any cases of wound infection from the available data.

## Discussion

Our results have demonstrated that, in this case series, at least 11.5% of CS were considered inappropriate by the expert panel. Considering that poor quality of records precluded the panel coming to a conclusion in 22%, and in a further 20% it was considered that alternative measures such as augmentation of labour or reassessment may have led to a different decision, it is possible that as many as half of the CS performed may have been avoided with different care. Given the risks of CS to both women and babies, it is important to consider why these potentially unnecessary surgical births are occurring. The process of making a decision to operate is complex with many contributory issues relating to capacity, human factors and both demand and supply-side concerns.

There was considerable variation in midwifery caseloads between the study facilities but all were high, indicating that midwives were under considerable pressure when providing care on the labour ward. For comparison, an annual midwifery caseload of 1 midwife to 28 patients has been reported in UK [11]. The pressure felt by midwives to get women delivered and off the labour ward may result in midwives referring women to doctors at an earlier stage than they otherwise would, had the staffing ratio been more favourable. Midwives may fear that they lack the capacity to provide the extra monitoring necessary when augmenting a labour with oxytocin, for example. Once a patient has been sent to theatre, midwives may feel better able to provide care to the women remaining on the labour ward. This may result in midwives making a case for CS to doctors when other measures could have resulted in a vaginal birth.

It has been suggested that doctors may be concerned about the possibility of litigation if the outcome is sub-optimal [12]. There may be a perception that by performing a CS they will be seen to have done everything that could be done. This concern, coupled with inadequate fetal heart rate monitoring, a lack of training about the significance of changes in the fetal heart rate and pattern and the presence of meconium in the liquor, may result in over-zealous diagnosis of suspected fetal hypoxia, commonly termed “non-reassuring fetal status”. We were not able to conduct key-informant interviews within the scope of this study to confirm these ideas, but the Kenyan expert assessors were of the view that fear of litigation and lack of training were both significant factors influencing the decisions of medical officers.

The staffing situation is frequently compounded by the redeployment of both midwifery and medical staff to other departments. Shikuku et [13] determined that, in the same counties as this study, 42.5% of health service staff who had undertaken emergency obstetric skills training over a five-year period were redeployed from the maternity unit. This frequently results in less experienced staff being placed on labour wards with the result of an erosion of decision-making skills.

The assessors failed to reach agreement concerning cases where the CS was performed due to the observed passage of fetal meconium in utero. Whereas in some cases the passage of meconium may occur in relation to gut hypoxia, the majority babies passing meconium in utero are born with no signs of hypoxia. Because of the risk of meconium aspiration in utero, if a fetus subsequently develops hypoxia after passing meconium, guidelines recommend that closer fetal monitoring should occur in such cases [14]. Some of the assessors felt that CS was justified in these cases because the labour ward staff lacked the capacity or equipment to escalate fetal monitoring appropriately. Whilst this may avoid some cases of meconium aspiration, undoubtedly it tends to increase the number of potentially avoidable CS. In this case series, the passage of meconium was documented in 28 (32%) of cases. Of these, 24 (85.7%) babies had 5-minute Apgar scores greater than 7. Of the four cases with meconium liquor and low Apgar scores, two had prolonged decision to delivery intervals, during which no fetal monitoring was recorded.

The WHO has recommended that a second opinion is sought before a decision to perform a CS is finalised [9], but in this study most decisions were made by non-specialist doctors without recourse to specialist opinion. In some situations, staffing constraints and lack of available consultants may have precluded this, but it is also possible that the prevailing clinical culture is such that doctors feel they may be regarded as incapable if they fail to make decisions on their own, despite a lack of training in CS decision-making.

In high income countries there has been much debate regarding the role of women’s choice in CS birth. For example, a study in Australia, found that 77% of obstetricians would agree to a CS on the grounds of maternal request [15]. In this study however, maternal request was only documented as a factor in one case. All the facilities included in this study were public facilities, and the proportion of women requesting and obtaining CS may be higher in private hospitals [16].

Prolonged decision to delivery intervals may contribute to both maternal and neonatal morbidity. In this study only four of the 87 cases were delivered in less than an hour of the decision for CS. Similar findings were reported in a study from Uganda based in the national referral hospital, where the median decision to delivery interval was 5.5 hours [17] and this was often related to the time of day the decision was made. In our study it was rare to find that the reasons for delay were documented, although in some cases this was stated to be due to the operating theatre being occupied by another case. Some facilities had a dedicated maternity theatre, whilst others shared operating facilities and staff with general surgeons. Clearly the performance of a non-indicated CS can have a knock-on effect resulting in delays to other cases, regardless of indication. In this study we found that once a decision for a CS had been made, in the majority of cases, there was no further documentation regarding fetal or maternal monitoring. Given that most women waited for 2 or more hours for their surgery this lack of monitoring is concerning. The fetal outcome in one of these cases was a fresh stillbirth.

### Strengths and limitations

This study utilised the experience and knowledge of the local context of the panel of expert reviewers. Each case was examined in depth by two reviewers and then discussed to ensure agreement, which was reached by the panel in all but five cases. To the best of our knowledge, this is the first such study using this methodology to review decisions for CS in the first stage of labour in Kenya to be published.

Extracted data was limited because of the poor quality of some of the records, and this constrained the ability of the team to determine the appropriateness of the CS in 21.8% of cases. The sample size in the study was relatively small due to limited funding and the time constraints of the reviewers taking part. This may limit the generalisability of the findings

## Conclusions

This study has demonstrated that there is room for improvement in CS decision-making in referral hospitals in Kenya. Whilst CS may seem a straightforward solution to an immediate concern, education of doctors and midwives should include raising of awareness regarding both short and long-term consequences, resulting in more nuanced decision-making. The quality of care provided on the labour ward could be improved by increases in staff to patient ratios, allowing staff to have adequate time for fetal and maternal monitoring, both during labour and after a decision for a CS birth has been made. The need to improve fetal monitoring may also be addressed by technical aids such as hand held doppler devices for intermittent auscultation or Cardiotocography, but appropriate training should be built into any such implementation. Interventions to provide in-service education should also include multi-disciplinary training on the required standards of record-keeping. The creation of tools such as customised stamps and proformas for recording examination findings may result in further improvements [17] and facilities should instigate a regular audit of CS indications to monitor trends in CS.

Ensuring that CS are performed for appropriate indications requires multiple interventions and investment. Developing a context-specific understanding as to the causes of inappropriate surgery is key to targeting such interventions. Further studies are indicated to understand whether these findings are representative of the wider situation in Kenya and beyond.

## Data Availability

All data produced in the present study are available upon reasonable request to the authors

## Acknowledgements

The authors gratefully acknowledge Lucy Nyaga and the members of the LSTM Kenya and UK teams for their support with this study, and Dr Paul Nyongesa for assistance with obtaining ethical approval from Moi University Institutional Research Ethics Committee.

## Funding

The study was partly funded under a grant from the UK FCDO (Reducing Maternal & Neonatal Death in Kenya, number 202549) (Charles Ameh), and partly from an award from LSTM (Fiona Dickinson). The funders had no role in study design, data collection and analysis, decision to publish, or preparation of the manuscript.

## Author contributions

Helen Allott: Conceptualisation, methodology, investigation, formal analysis, writing-original draft preparation & editing

Fiona Dickinson: Conceptualisation, funding acquisition, methodology, investigation, formal analysis, writing-review and editing

Stephen Karangau: Data interpretation, formal analysis, writing-review & editing

Michael Odour: Data interpretation, formal analysis, writing-review & editing

Nassir Shaban: Data interpretation, formal analysis, writing-review & editing

Evans Ogoti: Data interpretation, formal analysis, writing-review & editing

Sheila Sawe: Data interpretation, formal analysis, writing-review & editing

Ephraim Ochola: Data interpretation, formal analysis, writing-review & editing

Charles Ameh: Conceptualisation, funding acquisition, methodology, supervision, writing-review & editing

## Conflict of Interests

The authors have no conflict of interests

## Notes

### Competing Interest Statement

The authors have declared no competing interest.

